# Pseudotemporal ordering of spatial lymphoid tissue microenvironment profiles trails Unclassified DLBCL at the periphery of the follicle

**DOI:** 10.1101/2023.04.05.23288191

**Authors:** Claudio Tripodo, Giorgio Bertolazzi, Valeria Cancila, Gaia Morello, Emilio Iannitto

**Affiliations:** Tumor Immunology Unit, University of Palermo School of Medicine, Palermo, Italy; Histopathology Unit, The FIRC Institute of Molecular Oncology (IFOM), Milan, Italy; Department of Statistical Sciences, University of Palermo, Palermo, Italy; Department of Oncology, Hematology and Bone Marrow Transplants Unit La Maddalena, Palermo, Italy

**Keywords:** Digital spatial profiling, lymphoid tissue microenvironment, pseudotemporal ordering, Diffuse Large B cell Lymphoma, Cell-of-origin

## Abstract

We have established a pseudotemporal ordering for the transcriptional signatures of distinct microregions within reactive lymphoid tissues, namely germinal center dark zones (DZ), germinal center light zones (LZ), and peri-follicular areas (Peri). By utilizing this pseudotime trajectory derived from the functional microenvironments of DZ, LZ, and Peri, we have ordered the transcriptomes of Diffuse Large B-cell Lymphoma cases. The apex of the resulting pseudotemporal trajectory, which is characterized by enrichment of molecular programs fronted by TNFR signaling and inhibitory immune checkpoint overexpression, intercepts a discrete peri-follicular biology. This observation is associated with DLBCL cases that are enriched in the Unclassified/type-3 COO category, raising questions about the potential extra-GC microenvironment imprint of this peculiar group of cases. This report offers a thought-provoking perspective on the relationship between transcriptional profiling of functional lymphoid tissue microenvironments and the evolving concept of the cell of origin in Diffuse Large B-cell Lymphomas.

---

Diffuse large B-cell lymphomas (DLBCL) are phenotypically and genetically heterogeneous. Applying a classification algorithm to the gene expression profile of DLBCLs segregates the cases into two major subgroups^1^. Germinal Center B Cell-like (GCB) DLBCL subgroup shows a high level of expression of genes characteristic of physiological germinal center (GC) reaction molecular programs. Another DLBCL subgroup labelled Activated B Cell-like (ABC) expresses genes typical of mitogenically-activated B-cells, is enriched in plasma cell-related programs, and regarded as embodying post-GC dynamics. A third subgroup of DLBCL cases is left out from the GCB/ABC dichotomy, which does not express either set of genes at a high level and is accordingly named Unclassified or Type-3^2^. Based on the transcriptional profile similarity, GCB and ABC DLBCL are considered frozen in or stemming from a cell of origin (COO) at a different stage of the functional modulation pathway engaging B-cells in their GC journey towards Plasma cells or memory B^3^. At difference, the Unclassified/Type-3 DLBCL group has been considered as possibly consisting of more than one type of DLBCL and populated mainly by borderline cases not assigned by the clustering algorithm, yet actually belonging to the ABC or GCB group^2^. Several subsequent studies, applying different profiling platforms and classification algorithms, reproduced this molecular DLBCL tripartition, with the GCB representing the largest group (46-58%), followed by ABC (27-40%) and Unclassified/Type-3 (10-22%)^4–7^. Retrospective studies reported that GCB DLBCL show a significantly better overall survival (OS) curve than ABC; conversely, the Unclassified/Type-3 group outcome varies widely among different retrospective series, showing an OS curve comparable to that of ABC or GCB DLBCL or lying in between^4-7^. Of note, in two large prospective series, each including over a thousand patients, the Unclassified/Type-3 COO DLBCLs showed an outcome comparable to that of the ABC group^8,9^. Recently, the genetic heterogeneity and complexity underlying the COO expression profile have been partially elucidated. Comprehensive genomic analysis with different platforms led to the genetic subtype classification of DLBCLs^6,10,11^. With the caveat that nearly half of cases remained unclassified^6,12^, the genetic subtype classification of DLBCL highlighted that each of the three DLBCL COO gene expression subgroups included multiple genetic profiles. In particular, the Unclassified/Type-3 COO came out to be enriched for the BN2 genetic subtype that accounted for over one-third of cases, and comprised also cases belonging to ST2, EZB, MCD, A53 genetic clusters^12^. BN2 DLBCL are characterized by mutations that activate NOTCH2 or inactivate the NOTCH antagonist SPEN, frequently co-occurring with BCL6 translocations. Genetic aberrations targeting regulators of the NF-κB pathway are another prominent feature of BN2 DLBCL. Mutations targeting components of the BCR-dependent NF-kB pathway (PRKCB, BCL10, TNFAIP3, TNIP1) occur in over 50% of cases predicting that these tumours rely on B-cell receptor-dependent NF-κB activation and could be vulnerable to antagonists of B-cell receptor signalling^6^. The genomic profile of the BN2 cluster closely reminds that of Marginal Zone Lymphoma (MZL) and transformed MZL. All BN2 cases were confirmed to display a canonical DLBCL histological picture, suggesting that the Unclassified/Type-3 COO, besides misclassified BCL and GCB cases, may host a distinct subset of DLBCL^6^. However, whether the Unclassified/Type-3 group is merely the wastebasket of gene expression profile (GEP) classification algorithms or the profile of DLBCL originating from discrete functional differentiation stages and microenvironmental settings has not been elucidated.

We have exploited the spatial transcriptional profiling (1824 genes, cancer transcriptome atlas panel) of 15 microregions (Regions of Interest, ROIs) relative to GC dark zone (DZ, n=5) and light zone (LZ, n=5) microenvironment and peri-follicular (Peri n=5) areas. We used a pseudotime algorithm called PhenoPath^13^ to learn about the biological progression that characterizes the ROIs of interest. In the context of single-cell analysis, pseudo time is a computational construct that is used to order cells along a temporal trajectory, based on the similarity of their gene expression profiles. Pseudo time analysis aims to capture the temporal ordering of cells based on the expression changes of key genes, without directly measuring the time point at which the cells were collected. Pseudo time analysis can help to reveal the cellular processes that are active during different stages of development or differentiation, and can provide insights into the regulatory networks that govern these processes. The recent development of a pseudotime algorithm calibrated for bulk RNA-seq data^13^ allowed us to extract the latent temporal information from the digital spatial profiling ROIs. Using PhenoPath we estimated the pseudotemporal values from the bulk gene-expression matrix (supplementary table 1). Based on the pseudotime estimation, we obtained a pseudotemporal ordering of the individual ROIs. The pseudotime trajectory extended from DZ towards Peri regions, showing a clear association between pseudotemporal order and the spatial and functional ROI features (Figure 1A). Pseudotemporal ordering of digital spatial profiling ROIs enabled the identification of a pseudotime-associated gene signature composed of 184 genes significantly correlated with pseudotime (68 positively and 116 negatively) (Figure 1B, Supplementary Table 1). The genes positively associated with pseudotime were mostly enriched in TNF signaling pathway genes (i.e. *NFRSF14, TNFRSF25, TNFRSF1A, TNFRSF1B*) and in genes involved in the negative regulation of T-cell activation (i.e. *CD274, VSIR, LAG3, IDO1* – Supplementary Table 2) while those inversely associated with pseudotime were enriched in cell proliferation, DNA damage repair, and B-cell receptor signaling programs (Supplementary Table 2-3). In the attempt to investigate the effects of the pseudotime trajectory derived from the transcriptional profiling of functional microenvironments of a reactive lymphoid tissue on the ordering of DLBCL transcriptomes, we applied the 184-genes pseudotime signature to four independent gene expression-profiled DLBCL cohorts^4-7^ (Figure 2A-D). The four cohorts characterized by different case selection criteria and gene expression profiling technologies (Illumina, Affymetrix), displayed a generally conserved significance of the GCB vs ABC comparison in terms of OS (Supplementary Figure 1A-D). At odds, the fractions and prognostic behavior of Unclassified/Type-3 COO clusters in the four series were quite different (Supplementary Figure 1A-D), suggesting that this group of DLBCL might encompass a remarkable biological heterogeneity. By applying the pseudotime-associated microenvironment signature according to the tertile distribution of the cumulative expression of genes negatively and positively associated with pseudotime, DLBCL cases of the four cohorts displayed a common trend towards the enrichment of GCB cases in the *low-pseudotime* tertile (Figure 2E) a rather heterogeneous distribution of ABC cases, and a clear enrichment of Unclassified/Type-3 COO cases in the *high-pseudotime* tertile (Figure 2E). Consistently, the distribution of DLBCL genetic subgroups according to Schmitz across the low-, intermediate-, and high-pseudotime tertiles showed significant enrichment of EZB and other GCB genetics in the low-pseudotime group (Figure 2F, Supplementary Table 4). At the same time, BN2 was slightly enriched in the intermediate-pseudotime group, and other Unclassified genetics were detected across the three pseudotime categories, further indicating their mirroring of divergent biologies. We further explored if the pseudotime tertile hierarchy could rank DLBCL in groups with different prognosis. This hypothesis was tested in the series by Sha et al. (GSE117556)^7^, which offers distinctive features that are relevant to this extent: 1) a large prospective clinical data-set of 928 18-years or older DLBCL patients, with a centralized gene expression profiling and pathological review, eligible for anthracycline-based treatment; 2) 30 months Progression-Free Survival (PFS) in line with the best results of the recent phase 3 trials on DLBCL; 3) COO classification refined retrospectively with the same method, taking advantage of higher quality samples and improved data normalization over the complete data-set^7^. Most importantly, in this large series, the COO classification failed to identify groups with significantly different prognosis (Supplementary Figure 1E-F). The application of trichotomization of the series according to pseudotemporal scoring revealed significant prognostic differences between pseudotime-low and -high tertiles, with cases displaying high pseudotime ordering faring significantly better in terms of OS and PFS (Figure 2G, Supplementary Table 5). The different prognostic performance of COO and pseudotemporal ordering in this setting suggests that the transcriptional modulations represented in the spatial profiling of diverse GC and extra-follicular microregions may adequately cope with the wide continuum of DLBCL.

**Figure 1:**
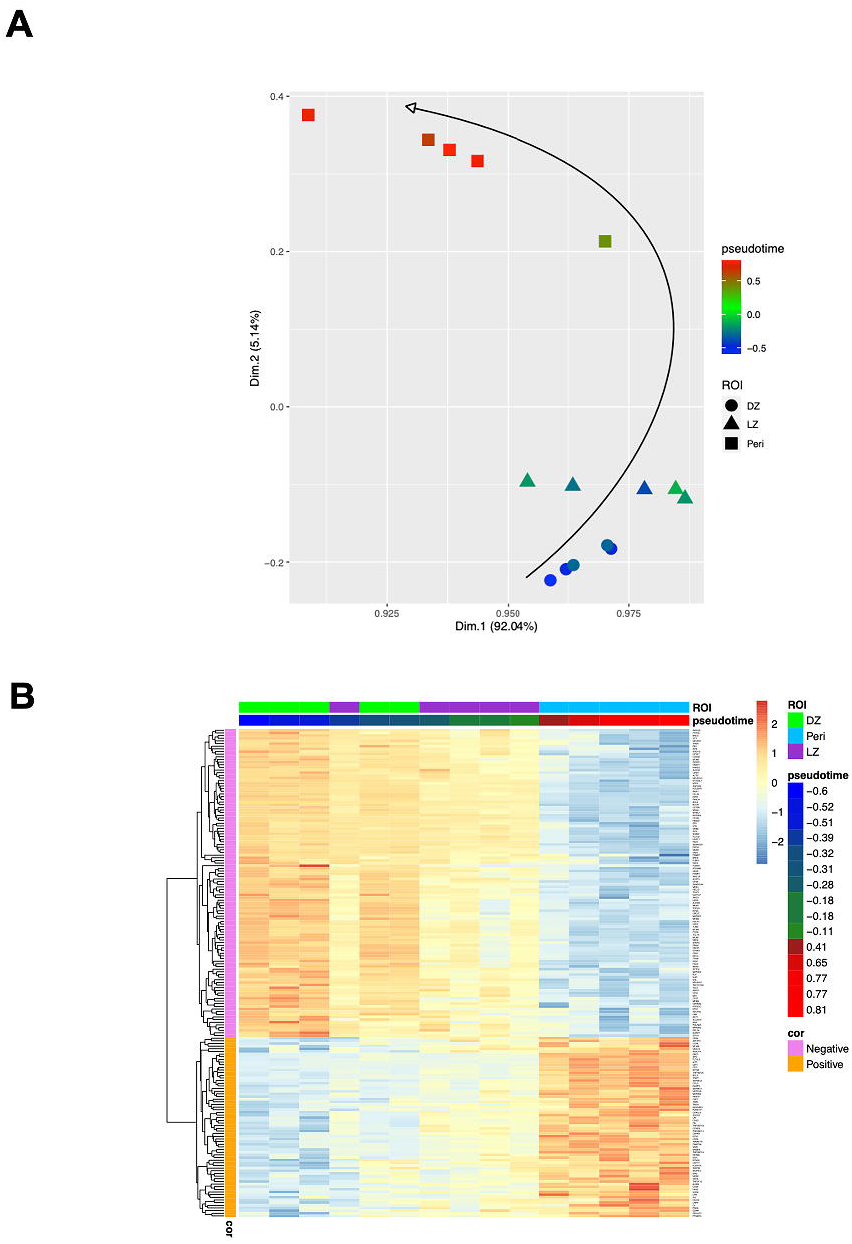
**A**, Two-dimensional principal component projection produces a trajectory among DZ, LZ, and Peri ROIs. The color gradient of points reflects the pseudotime estimated values. **B**, Expression heatmap of the 184 genes significantly correlated with the pseudotime over 15 ROIs. The 15 ROIs (columns) are ordered according to the pseudotime estimation.

**Figure 2:**
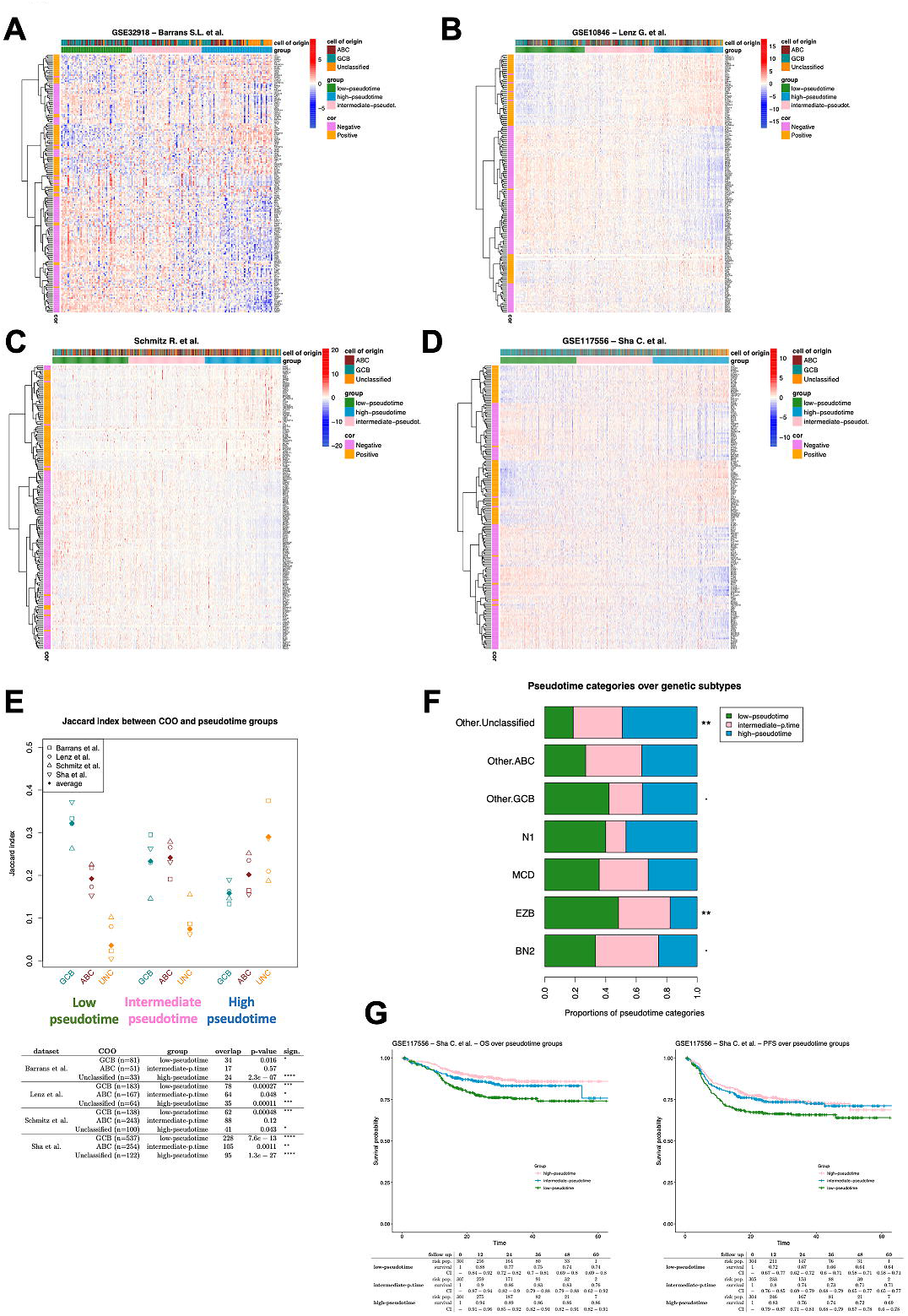
**A-D**, Expression heatmap of the 184 pseudotime-related genes. The DLBCL cases (columns) are ordered according to their pseudotime-related score (see methods). Three pseudotime groups have been identified by applying the tertile separation on pseudotime-related scores (i.e., low, intermediate, and high pseudotime groups). **E**, Jaccard similarity index between COO and pseudotime groups over DLBCL datasets. Unclassified cases strongly enrich all the high-pseudotime groups. While GCB cases enrich all the low-pseudotime groups (Fisher p-values are shown in the table). **F**, Proportions of pseudotime categories over genetic subtype groups. The Fisher exact test has been applied to evaluate the association between genetic subtypes and pseudotime categories (Suppl. Table 4). **G**, Survival analysis on DLBCL cases from Sha et al. dataset. Patients were divided into three groups according to the tertiles of their pseudotime-related scores (i.e., low, intermediate, and high pseudotime groups).

Our results also demonstrate that the apex of the pseudotemporal trajectory resulting from spatial profiling intercepts a discrete peri-follicular biology characterized by enrichment of molecular programs fronted by TNFR signaling and inhibitory immune checkpoint overexpression and corresponding to cases enriched in the Unclassified/type-3 COO category, opening an issue regarding the potential extra-GC imprint of this heterogeneous group of cases. Moreover, an accurate biomolecular characterization of this hitherto neglected subset of DLBCL might pave the way for deciphering their biological and prognostic determinants.

## Data Availability

All data produced in the present work are contained in the manuscript

https://onlinelibrary.wiley.com/doi/full/10.1002/eji.202149746

https://www.ncbi.nlm.nih.gov/geo/query/acc.cgi?acc=GSE32918

https://www.ncbi.nlm.nih.gov/geo/query/acc.cgi?acc=GSE10846

https://www.ncbi.nlm.nih.gov/geo/query/acc.cgi?acc=GSE117556

## Notes

The Authors have no conflict of interest to disclose

### Competing Interest Statement

The authors have declared no competing interest.

### Funding Statement

This study was funded by the Italian Foundation for Cancer Research (AIRC)

### Author Declarations

All the datasets used in the study are publicly available at the following links:: https://onlinelibrary.wiley.com/doi/full/10.1002/eji.202149746 https://www.ncbi.nlm.nih.gov/geo/query/acc.cgi?acc=GSE32918 https://www.ncbi.nlm.nih.gov/geo/query/acc.cgi?acc=GSE10846 https://www.ncbi.nlm.nih.gov/geo/query/acc.cgi?acc=GSE117556

