## Supplementary figures and images for "Pseudotemporal ordering of spatial lymphoid tissue microenvironment profiles trails Unclassified DLBCL at the periphery of the follicle"

### Supplementary Figure 1

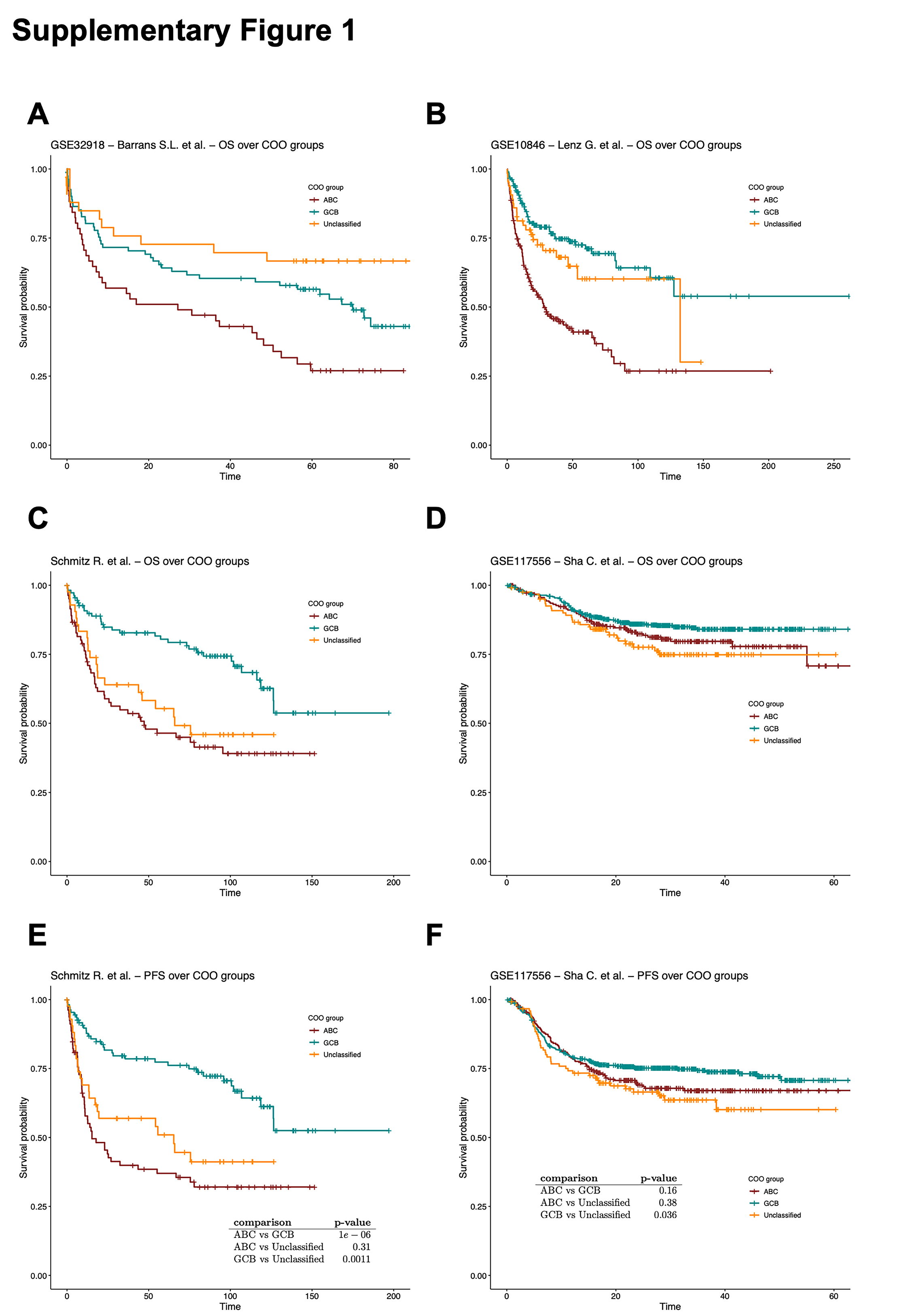
