## Supplementary Methods for "Pseudotemporal ordering of spatial lymphoid tissue microenvironment profiles trails Unclassified DLBCL at the periphery of the follicle"

*Digital spatial profiling*

As described in our previous work14, the transcriptional landscape of 15 different spatially-resolved regions of interest (ROIs) of the tonsil (5 peri/inter-follicular ROIs, 5 DZ, and 5 LZ ROIs from morphologically normal follicles) was determined by Digital Spatial Profiling on slides stained with CD271/NGFR (as an follicular dendritic cells marker to highlight the LZ) and CD20 (as a B-cell marker). The 15 selected and segmented ROIs were profiled using a GeoMx Digital Spatial Profiler (DSP) (NanoString, Seattle, WA) applying the Cancer Transcriptome Atlas panel (<https://www.nanostring.com/products/geomx-digital-spatial-profiler/geomx-rna-assays/geomx-cancer-transcriptome-atlas/>).

*Methods*

Raw counts were normalized against the 75th percentile of signal from their own ROI. ­The R package Phenopath [2] has been used to estimate pseudotime values from bulk gene expression data as described by Campbell and Yau [3]. We choose of the Phenopath algorithm for the pseudotime estimation because the standard pseudotime algorithms require single-cell data as input, while Phenopath can be used also on bulk RNA-seq data.

Therefore, we consider a pseudotemporal ranking of ROIs based on pseudotime estimations. The temporal trajectory has been highlighted on a PCA projection performed on normalized data using the FactoMine R package.

The Spearman correlation coefficients have been calculated between gene expression and pseudotime estimated values. The Bonferroni correction for multiple comparisons has been applied to evaluate the p-value significance of correlation coefficients (FWER controlled at 5% level). The pseudotime significantly correlated genes compose the pseudotemporal signature (Supplementary Table 1).

The pseudotemporal signature was assessed in the following DLBCL datasets: Barrans et al. (GSE32918) [4], Lenz et al. (GSE10846) [5], Schmitz et al. [6], and Sha et al. (GSE117556) [7]. The Barrans and the Sha datasets have been downloaded from GEO using the GEOquery R package. Regarding the datasets of Barrans, Sha, and Schmitz, we maintained the normalization proposed by the authors. The Lenz et al. (GSE10846) expression matrix has been obtained from the CEL file available on GEO and it has been normalized using the gcrma package.

To order DLBCL patients according to the pseudotemporal gene signature, we have calculated a pseudotime-related score that combines the expression of the pseudotime-signature genes with the correlation coefficients previously calculated on the DSP dataset. Considering the patient-*j*, his pseudotime-related score is calculated as:

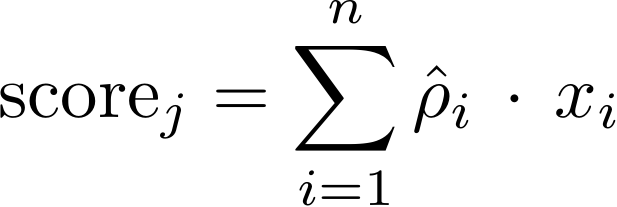

Where ρ_i_ is the Spearman correlation coefficient between the expression of gene-*i* and pseudotime values (it has been previously calculated on the DSP dataset), *x_i_* is the expression of gene-*i* in the DLBCL dataset, and *n* is the number of genes of the pseudotemporal signature. The coefficients ρ_i_ allow us to weight the gene expression considering how strong is the correlation between each gene and the pseudotime values. Using the score, each DLBCL cohort was divided into low-pseudotime, intermediate-pseudotime, and high-pseudotime.

The Jaccard similarity index has been calculated to measure the association between the cell of origin (COO) and the pseudotime groups in DLBCL datasets. The Fisher exact test has been used to evaluate the association between pseudotime groups and COO over DLBCLs, and the association between pseudotime groups and genetic subtypes in the Schmitz dataset.

The prognostic power of the pseudotemporal signature has been tested on the Sha et al. dataset. The overall Survival (OS) and the progression-free survival (PFS) have been compared among the three pseudotime groups. Kaplan-Meier method has been used to estimate the survival functions among groups, and the log-rank test has been used to test the differences in the overall survival between the identified groups. Before calculating the log-rank test, the cox-pzh test was used to test the proportional hazard assumption (Supplementary Tab.5). We have adapted a multivariate Cox model including the pseudotime groups, the COO classes, and the IPI-risk classes (i.e., low, medium, and high risk) to verify that the pseudotime group variable maintains its significance. The whole survival analysis has been carried out through the survival R package. All statistical analyses were performed using R software (v 4.0.2) (http://www.R-project.org).

*References*

1. L’Imperio V, Morello G, Cancila V, Bertolazzi G, Mazzara S, Belmonte B, Balzarini P, Corral L, Di Napoli A, Facchetti F, Pagni F, Tripodo C. *In situ* transcriptional profile of a germinotropic plasmablastic burst hints at an unfavorable DLBCL subset.
2. Campbell K (2022). phenopath: Genomic trajectories with heterogeneous genetic and environmental backgrounds. R package version 1.22.0.
3. Campbell KR and Yau C. Uncovering pseudotemporal trajectories with covariates from single cell and bulk expression data. Nature Communications volume 9, Article number: 2442 (2018)
4. Barrans SL, Crouch S, Care MA, Worrillow L, Smith A, Patmore R, Westhead DR, Tooze R, Roman E, Jack AS. Whole genome expression profiling based on paraffin embedded tissue can be used to classify diffuse large B-cell lymphoma and predict clinical outcome. Br J Haematol. 2012 Nov;159(4):441-53. doi: 10.1111/bjh.12045. Epub 2012 Sep 13. PMID: 22970711.
5. Lenz G, Wright G, Dave SS, Xiao W, et al. Stromal gene signatures in large-B-cell lymphomas. N Engl J Med 2008 Nov 27;359(22):2313-23.
6. R. Schmitz, L.M. Staudt et al. Genetics and Pathogenesis of Diffuse Large B-Cell Lymphoma. n engl j med 378;15 nejm.org April 12, 2018.
7. Sha C, Barrans S, Cucco F, Bentley MA, Care MA, Cummin T, Kennedy H, Thompson JS, Uddin R, Worrillow L, Chalkley R, van Hoppe M, Ahmed S, Maishman T, Caddy J, Schuh A, Mamot C, Burton C, Tooze R, Davies A, Du MQ, Johnson PWM, Westhead DR. Molecular High-Grade B-Cell Lymphoma: Defining a Poor-Risk Group That Requires Different Approaches to Therapy. J Clin Oncol. 2019 Jan 20;37(3):202-212. doi: 10.1200/JCO.18.01314. Epub 2018 Dec 3. Erratum in: J Clin Oncol. 2019 Apr 20;37(12):1035. PMID: 30523719; PMCID: PMC6338391.
